# Mediators of monocyte chemotaxis and matrix remodeling are associated with the development of fibrosis in patients with COVID-19

**DOI:** 10.1101/2023.04.28.23289261

**Authors:** Sarah E. Holton, Mallorie Mitchem, Sudhakar Pipavath, Eric D. Morrell, Pavan K. Bhatraju, Jessica A. Hamerman, Cate Speake, Uma Malhotra, Mark M. Wurfel, Steven Ziegler, Carmen Mikacenic

## Abstract

Acute respiratory distress syndrome (ARDS) has a fibroproliferative phase that may be followed by pulmonary fibrosis. This has been described in patients with COVID-19 pneumonia, but the underlying mechanisms have not been completely defined. We hypothesized that protein mediators of tissue remodeling and monocyte chemotaxis are elevated in the plasma and endotracheal aspirates of critically ill patients with COVID-19 who subsequently develop radiographic fibrosis.

We enrolled COVID-19 patients admitted to the ICU who had hypoxemic respiratory failure, were hospitalized and alive for at least 10 days, and had chest imaging done during hospitalization (*n* = 119). Plasma was collected within 24h of ICU admission and at 7d. In mechanically ventilated patients, endotracheal aspirates (ETA) were collected at 24h and 48-96h. Protein concentrations were measured by immunoassay. We tested for associations between protein concentrations and radiographic evidence of fibrosis using logistic regression adjusting for age, sex, and APACHE score.

We identified 39 patients (33%) with features of fibrosis. Within 24h of ICU admission, plasma proteins related to tissue remodeling (MMP-9, Amphiregulin) and monocyte chemotaxis (CCL-2/MCP-1, CCL-13/MCP-4) were associated with the subsequent development of fibrosis whereas markers of inflammation (IL-6, TNF-α) were not. After 1 week, plasma MMP-9 increased in patients without fibrosis. In ETAs, only CCL-2/MCP-1 was associated with fibrosis at the later timepoint.

This cohort study identifies proteins of tissue remodeling and monocyte recruitment that may identify early fibrotic remodeling following COVID-19. Measuring changes in these proteins over time may allow for early detection of fibrosis in patients with COVID-19.

## Introduction

The acute respiratory distress syndrome (ARDS) is a highly morbid, often fatal syndrome that affects approximately 23% of patients requiring mechanical ventilation^1^. Pulmonary fibrosis following fibroproliferative ARDS has been described previously^2,3^, and was reported following the SARS epidemic of 2003^4,5^ and MERS outbreak of 2012^6^. The COVID-19 pandemic has caused severe respiratory failure in many patients^7–10^, with longitudinal studies showing that a proportion of survivors of COVID-19 have ongoing respiratory symptoms^11^ and decreased lung function^12^, while others have persistent radiographic abnormalities including fibrosis^13–16^. Further investigation is needed to understand why this syndrome has such heterogeneous effects and to identify potential therapies that can be used during future viral pandemics. Patients with COVID-19 have a three-fold longer duration of mechanical ventilation compared to patients with influenza^17^, which may be due in part to the development of a fibroproliferative phase after viral pneumonia and respiratory failure in patients with severe disease^18–21^.

While an associative link between viral ARDS and fibrotic lung remodeling has been established, the underlying mechanism in humans remains unclear. Unlike other studies of post-ARDS fibrosis in which patients have had multiple etiologies for ARDS, studying patients with COVID-19 ARDS provides a unique opportunity to understand the timeline of lung injury and remodeling. Further, with growing numbers of survivors of COVID-19 we are beginning to understand the long-term sequelae both in the lungs and elsewhere in the body. Recent studies have identified pro-fibrotic macrophage populations in the lungs of patients with COVID-19, potentially from recruited circulating monocytes, consistent with prior studies showing monocyte recruitment is linked to the development of pulmonary fibrosis following injury^18,22^. Other groups have analyzed transcriptional changes in lungs from patients undergoing lung transplant or at autopsy and identified pro-fibrotic programs that are similar to those in idiopathic pulmonary fibrosis^19–21^. This important work establishes a link between fibrotic remodeling during fibroproliferative ARDS and other fibrotic lung diseases, including post-ARDS fibrosis.

Prior studies have established some proteins identified in the circulation or alveolar space that are associated with fibroproliferative ARDS^23–28^. We hypothesized that proteins previously associated with fibrotic lung disease, including idiopathic pulmonary fibrosis, would be elevated in blood and lung fluid from critically ill patients with SARS-CoV-2 who go on to develop fibroproliferative ARDS or other radiographic features of fibrosis. We measured these proteins in the plasma and endotracheal aspirates of patients early during their hospital course to determine which pathways may be involved in the pathogenesis of disease, prior to the development of radiographic fibrotic features.

## Results

### Patients in the ICU with COVID-19 developed radiographic features of fibrosis during hospitalization

Patients were enrolled from three University of Washington ICUs with suspected COVID-19 as previously described^29–31^ (*n* = 366) (**Figure 1**). To study the effects of COVID-19 on the development of radiographic features of fibrosis (hereafter referred to as “fibrosis”), only those patients with confirmed COVID-19 (based on a positive SARS-CoV-2 PCR test) were included. The chest x-ray or CT scan closest to discharge or death was used to classify fibrosis status. On chest x-rays, presence of severe traction bronchiectasis or radiologist report was used to determine fibrosis status (**Supplemental Figure 1**). On CT scans, reticulation, honeycombing, and traction bronchiectasis were quantified by a blinded chest radiologist (**Supplemental Table 1**). Examples of nonfibrotic and fibrotic chest CTs are shown in **Figure 2**.

**Figure 1.**
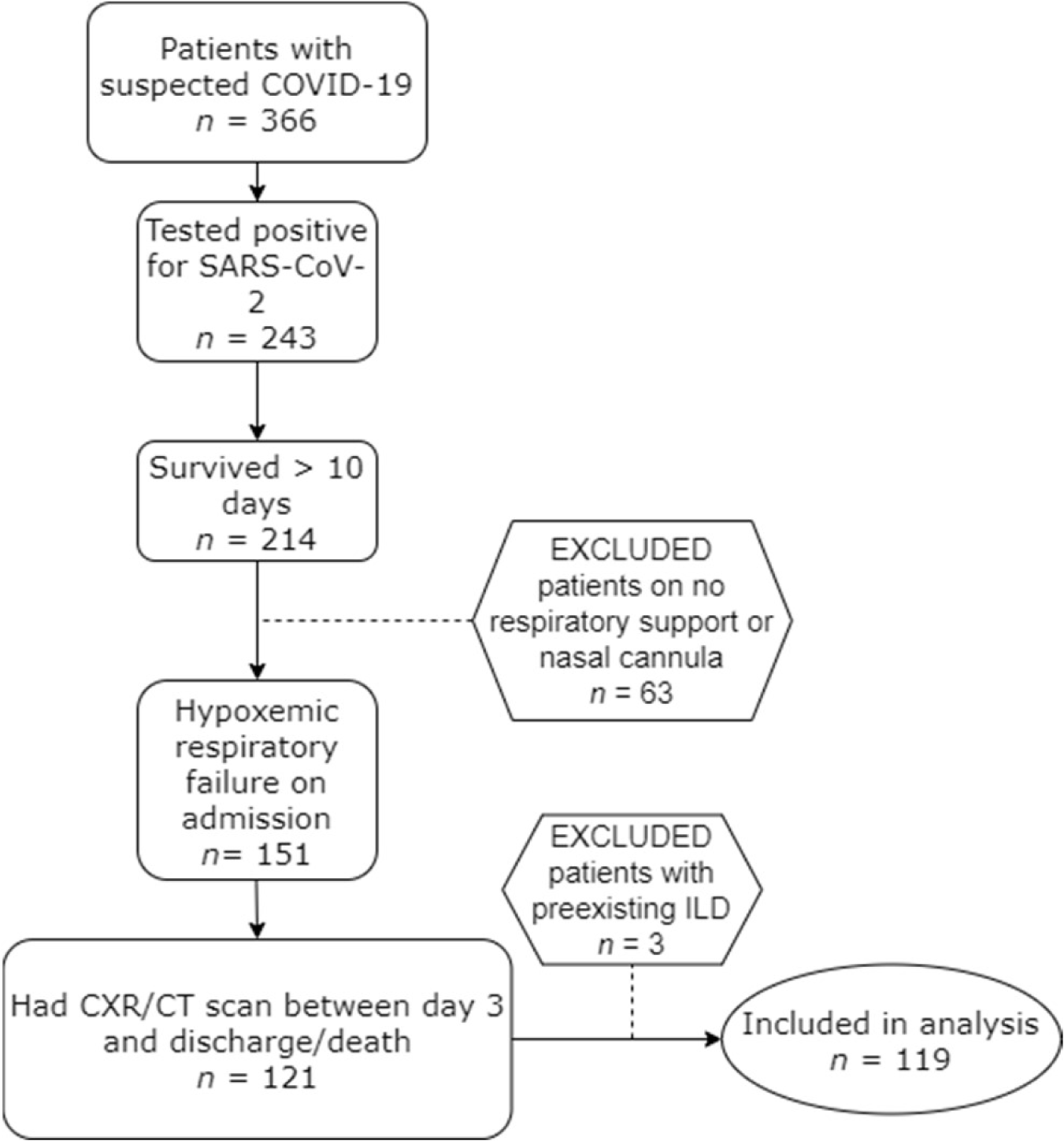
Development of cohort. 119 patients were included in biomarker analysis. Three patients had pre-existing interstitial lung disease (ILD) from sarcoidosis, idiopathic pulmonary fibrosis, and fibrotic hypersensitivity pneumonitis and were excluded from this analysis.

**Figure 2.**
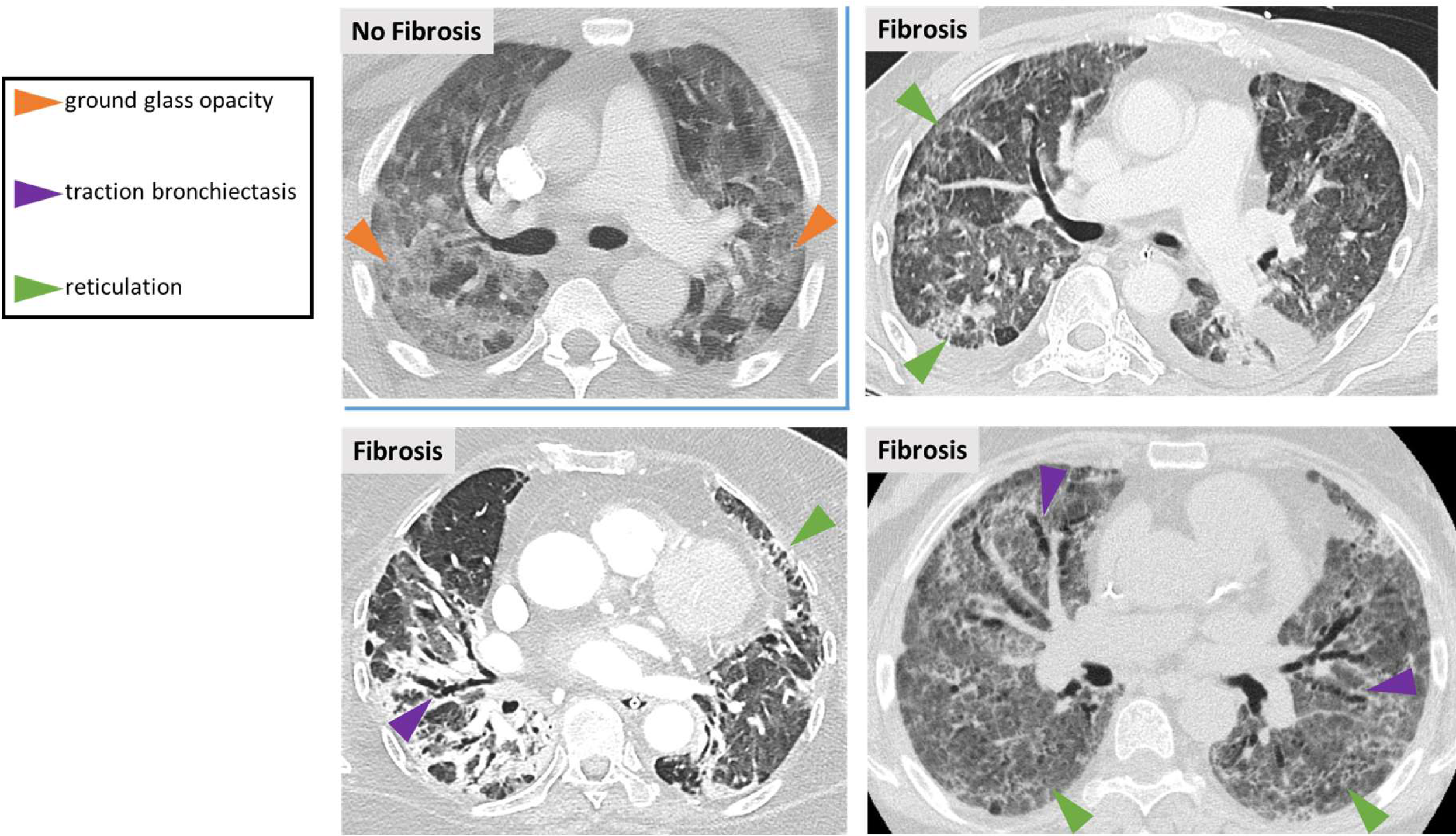
Evaluation of chest CTs for presence of fibrotic features. Fibrotic features were defined as reticulation, traction bronchiectasis, or honeycombing. Patients with predominantly ground glass opacities on chest CTs were categorized as “no fibrosis”.

We identified 39 patients with fibrosis and 80 patients with no evidence of fibrotic features. Of the patients with fibrosis identified on chest CT, 11/25 had >5% traction bronchiectasis (**Supplemental Table 1**). Demographics including age, race and ethnicity were similar between the two groups (**Table 1**). Patients with fibrosis tended to have higher APACHE scores but similar P:F ratios at the time of enrollment. Patients with fibrosis had a longer time interval between their qualifying positive SARS-CoV-2 PCR test and the first day of sampling and included a higher percentage of patients transferred from an outside hospital (82% compared to 57%) (**Table 1**). When we stratify by survival status, there was no difference between SARS-CoV-2 PCR test and enrollment among survivors (*p* = 0.16) (**Supplemental Figure 2a**).

**Table 1:**
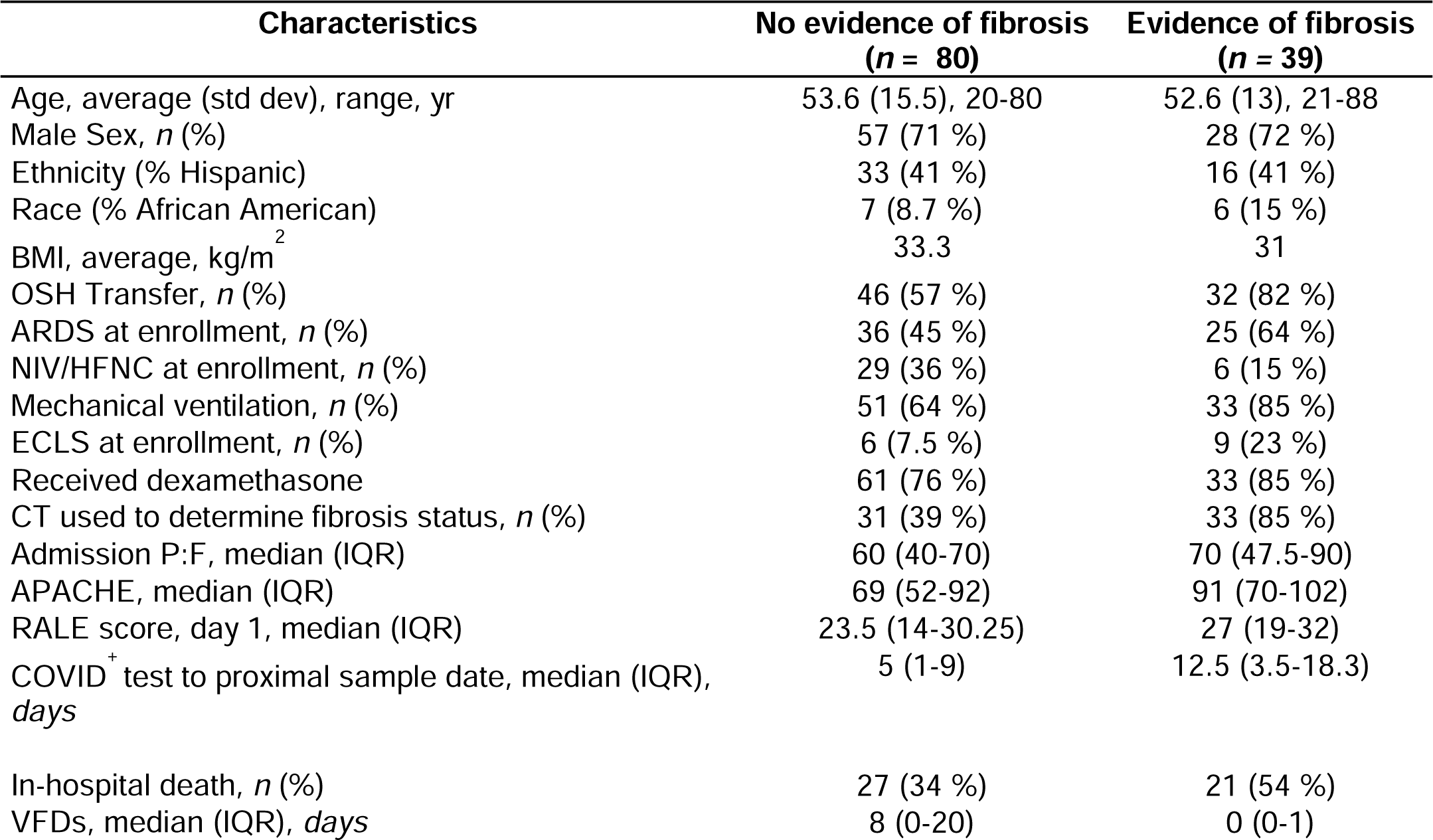
Clinical characteristics of patients within cohort (*n* = 119). In general, patients with radiographic features of fibrosis were more clinically ill at the time of enrollment than patients without fibrosis. This is reflected in the higher percentage that had ARDS by Berlin criteria at the time of enrollment, were on extracorporeal life support, and enrollment APACHE III scores. These patients also had fewer ventilator free days (VFDs) and higher in-hospital mortality.

Patients with fibrosis also had significantly longer hospital stays (**Supplemental Figure 2b**); this was true of survivors (*p* <0.0001) and non-survivors (*p* =0.0009). Patients with fibrosis had a longer time interval between enrollment and CXR/CT scan used to determine fibrosis status (*p* = 0.01 for nonsurvivors, *p* <0.0001 for survivors) though some survivors without fibrosis had imaging >50 days after COVID-19 diagnosis (**Supplemental Figure 2c**).

### Putative markers of pulmonary fibrosis are elevated in plasma early ICU course in patients with radiographic features of fibrosis

We measured markers of inflammation, monocyte chemotaxis, and matrix remodeling via immunoassay in our cohort in both plasma and endotracheal aspirates. Our primary hypothesis was that markers of monocyte chemotaxis and matrix remodeling would be elevated in patients with fibrosis. First, we analyzed markers of monocyte chemotaxis, CCL-2/MCP-1 and CCL-13/MCP-4 **(Figure 3a)** because higher numbers of circulating monocytes in the blood have previously been associated with pulmonary fibrosis^22^. We found that both CCL-2/MCP-1 (OR=1.45, [1.02-2.1], *p* = 0.038) and CCL-13/MCP-4 (OR = 2.6, [1.15-5.8], *p* = 0.02) were associated with higher risk of developing fibrosis. All associations with fibrosis are of log_2_-transformed biomarkers adjusted for age, sex, and enrollment APACHE score. CCL-13/MCP-4 has not previously been associated with pulmonary fibrosis or ARDS illness severity. Monocyte levels measured peripherally on routine complete blood count tests were no different between groups at the time of enrollment (**Supplemental Figure 3**).

**Figure 3.**
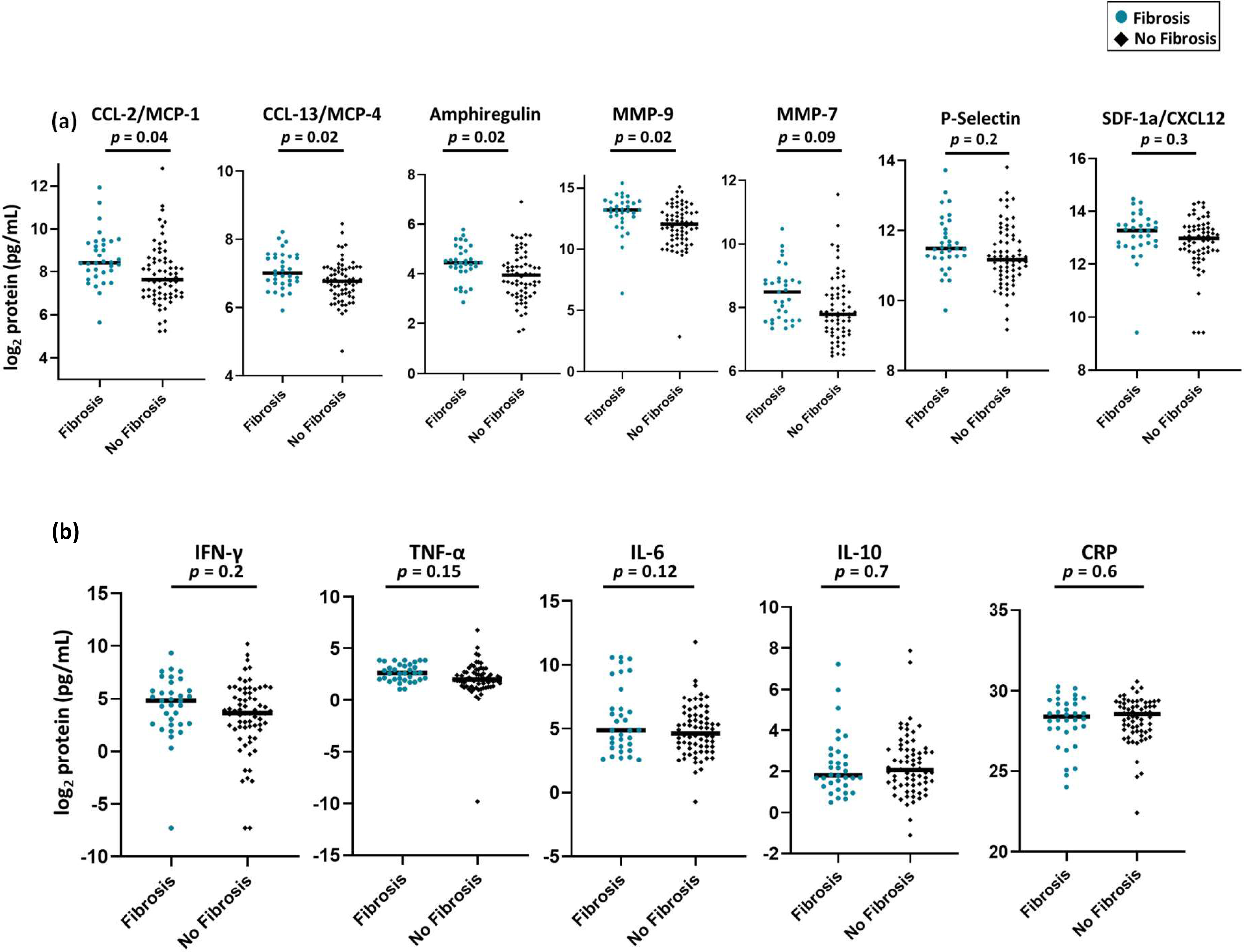
Markers of monocyte chemotaxis and matrix remodeling are higher in patients with radiographic features of fibrosis when measured in plasma within 24h of ICU admission. (a) Markers of monocyte chemotaxis, Amphiregulin, and MMP-9 are elevated in patients with fibrosis, whereas MMP-7, P-Selectin. and SDF-1a/CXCL12 are not. (b) IFN-γ and other markers of inflammation are not associated with fibrotic features. *p* values are derived from logistic regression with log_2_-transformed protein concentrations were adjusted for age, sex, and enrollment APACHE score

We then tested protein mediators of tissue remodeling that had been previously associated with pulmonary fibrosis. These included Amphiregulin, Matrix Metalloproteinase-7 (MMP-7), Matrix Metalloproteinase-9 (MMP-9), and Stromal Derived Factor-1alpha (SDF1a/CXCL12) ^32,33,34(p9),35–39^. We found that Amphiregulin (OR = 1.98 [1.1-3.5], *p*= 0.02) and MMP-9 (OR = 1.5, [1.1-2.1], *p*= 0.02) were associated with fibrosis while MMP-7 (OR = 1.5, [0.8-2.4], *p* =0.09), P-Selectin (OR = 1.4, [0.8-2.5], *p* = 0.2), and SDF-1a/CXCL12 (OR = 1.3, [0.8-2.3], *p* =0.3) were not at this early timepoint **(Figure 3a).**

Next, we determined whether markers of inflammation were different between patients with and without fibrosis (**Figure 3b**). IFN-γ, TNF-α, and IL-6, which have been suggested to reflect illness severity in COVID^40,41^, were not different between groups (OR 1.1, [0.95-1.3], *p* =0.2, OR=1.4 [0.9-2.1], *p* = 0.15, OR=1.2, [0.96-1.4], *p* = 0.12, respectively). C-reactive protein (OR = 0.9, [0.7-1.2], *p* = 0.4) and IL-10 (OR = 0.9, [0.7-1.3], *p* = 0.86) were also not different between groups. These data suggest that while our patients with fibrosis had higher indices of illness severity, inflammatory plasma proteins previously linked to a “cytokine storm” were not associated with development of fibrosis. When dexamethasone treatment was used as a co-variate in the regression model (**Supplemental Table 2**), there were no significant differences in the markers of monocyte chemotaxis or tissue remodeling, but IL-6 and TNF-α were also associated with fibrosis (OR 1.2, [1.0-1.5], p=0.03 and OR 1.6, [1.0-2.4], p=0.04, respectively).

A subset of patients had plasma sampled within 24h of study enrollment and again approximately 7 days later (*n* = 82/119). These patients were not pre-selected for multiple measurements and the lack of a serial measurement was not related to study drop-out. (Of the 18 patients without a 7d sample, 6 died, which is a similar mortality to our overall cohort. All patients included in the study were alive and remained in the ICU at day 10). When we looked at log_2_ transformed tissue remodeling markers (adjusted for age, sex, and enrollment APACHE score) at the 7 day timepoint, we found that only MMP-7 (OR = 1.6 [1.02-2.5], *p* =0.04) was associated with fibrosis (**Supplemental Table 2**). However, when we looked at how these markers changed over time **(Figure 4),** we found that MMP-7 levels increased over time in both groups of patients, and the fold-change between groups was not significant (FC = 1.16 [0.85-1.6], *p*= 0.34). This may reflect ongoing inflammation or remodeling that is not necessarily linked to the development of fibrosis. MMP-9, however, exhibited no change in patients with fibrosis whereas it increased significantly in patients without fibrosis (FC = 0.45 [0.3-0.7], *p* = 0.0017). There was no significant pattern of change in SDF-1a/CXCL12 (*p*=0.3), Amphiregulin (*p*=0.4), or P-Selectin (*p*=0.54).

**Figure 4.**
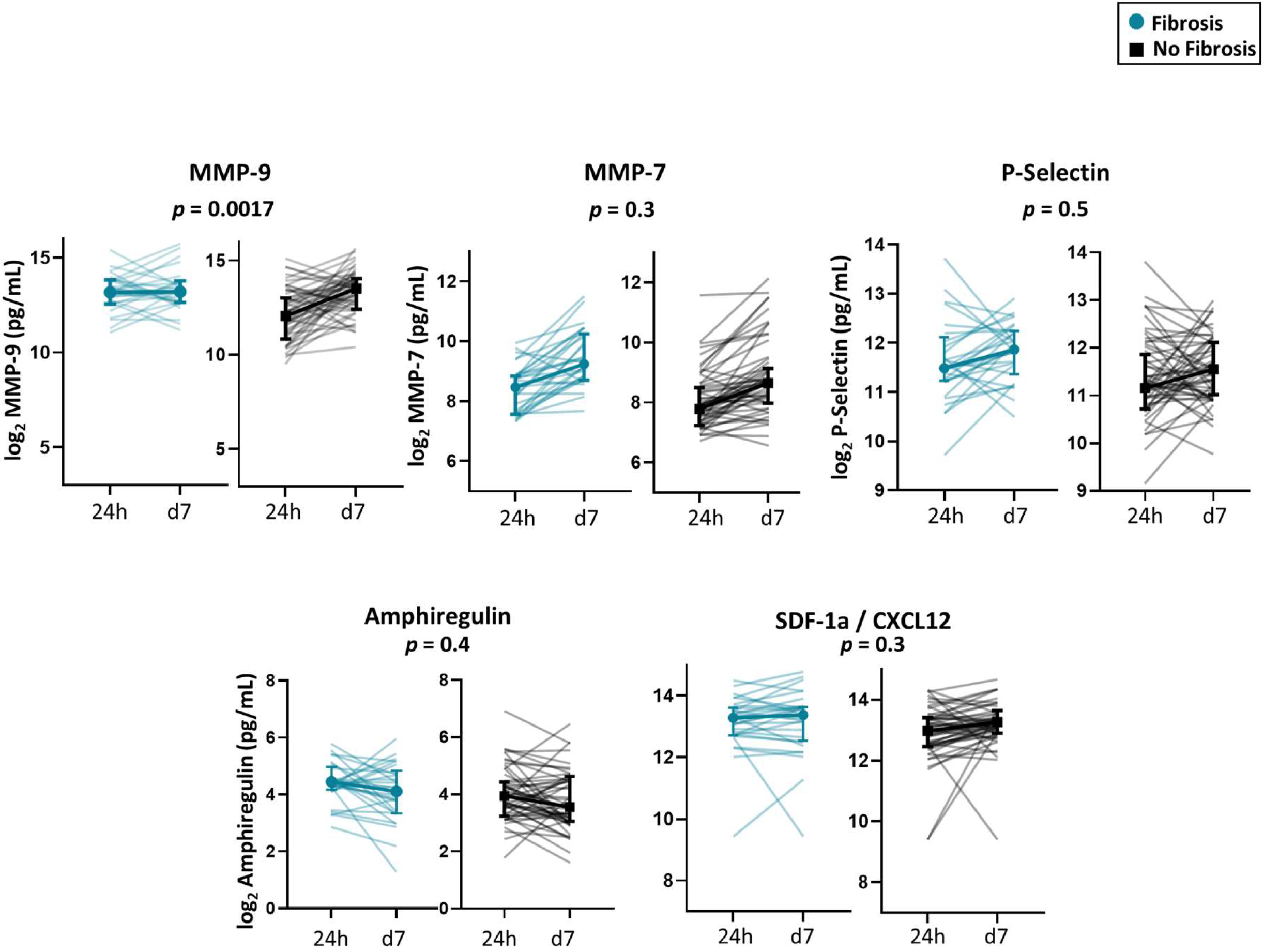
Plasma levels of tissue remodeling change over time. A subset of patients had markers measured both within 24h of ICU admission and again 7 days later. MMP-9 levels increased in patients without fibrosis and did not change in patients with fibrosis. MMP-7 levels increased in both groups of patients. Amphiregulin levels appeared to decrease over time in both groups of patients. P-Selectin and SDF-1a/CXCL12 levels did not have any consistent pattern in either group of patients. *p* values were calculated using linear regression of an association of the fold change of the later timepoint over the earlier timepoint and adjusted for age, sex, and APACHE score.

To ensure that potential misclassification of patients fibrosis status in those without chest CT scans did not affect our results, a sensitivity analysis was performed looking only at those patients with CT scans (**Supplemental Table 3**) and these results were not substantially different.

### Endotracheal aspirate measurements of tissue remodeling proteins are not associated with the development of fibrosis when measured at 24 and 72 hours after study enrollment

A subset of patients from our initial cohort study combined with patients enrolled in a separate ICU in Seattle, WA (combined *n* = 74, *n* = 27 with fibrosis) had endotracheal aspirate measurements collected within 24h of ICU admission and again approximately 72 hours later (**Supplemental Table 4**). The combined cohort had the same inclusion/exclusion criteria as the initial cohort, and sampling was done within the same time frame. All patients in the combined cohort were mechanically ventilated at the time of study enrollment (**Supplemental Table 5**). The chest radiologist did not review these chest images, and only official radiology reports were used to identify fibrosis/no fibrosis. The same markers were measured, although IFN-γ, IL-6, CCL-13/MCP-4, and SDF-1a did not meet quality control **(Supplemental Table 6).**

In models adjusted for age, sex, and hospital cohort, no individual marker was associated with fibrosis at the 24h timepoint, although CCL-2/MCP-1 trended towards higher concentrations in patients with fibrosis (OR = 1.3, [0.9-1.7], *p* =0.12) **(Figure 5a)**. Only CCL-2/MCP-1 was elevated and nearly associated with fibrosis (OR = 1.2, [0.99-1.5], *p* = 0.05) at the 48-96h timepoint **(Supplemental Figure 4).**

**Figure 5.**
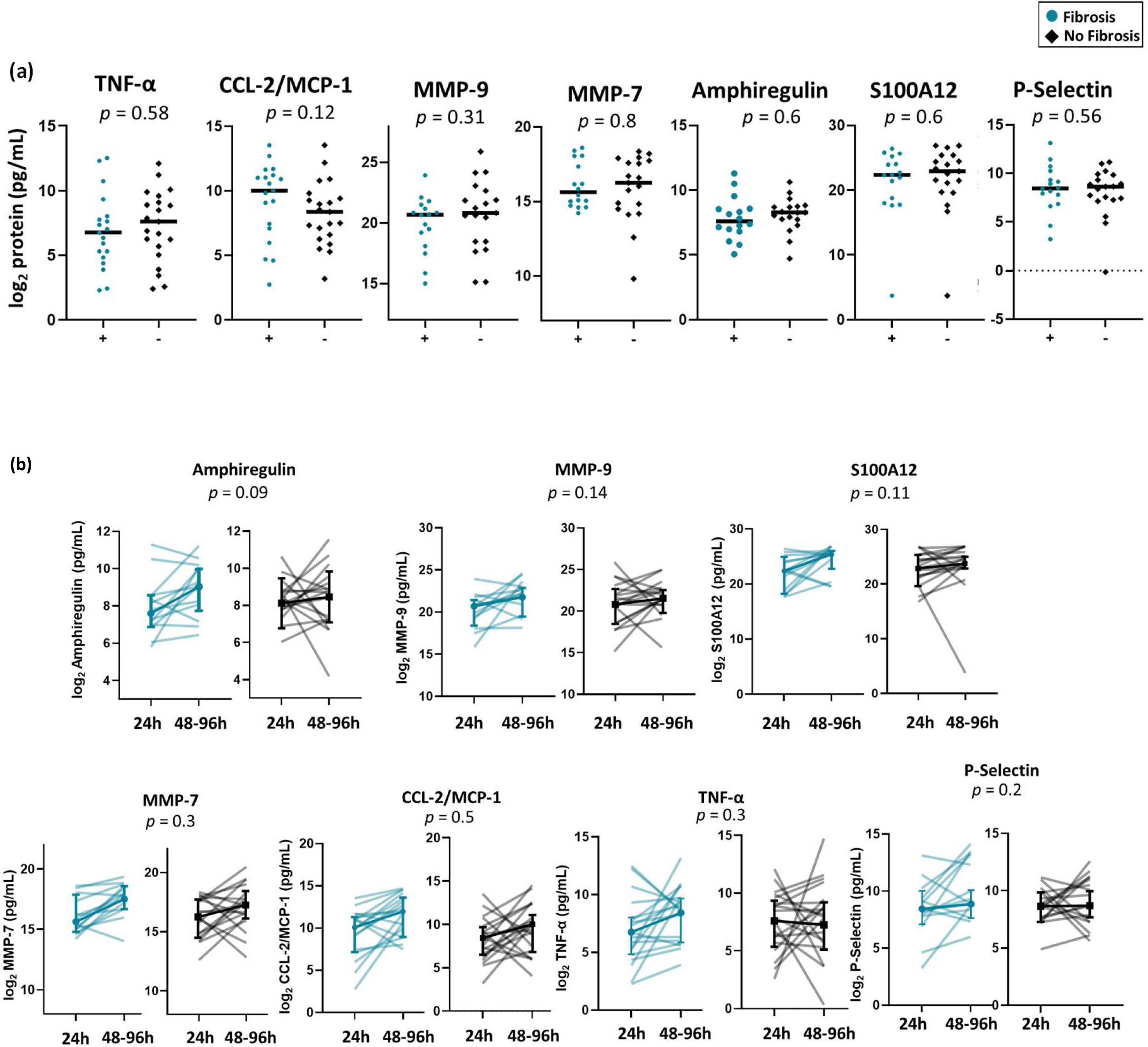
Protein levels measured in endotracheal aspirates at two timepoints are not associated with fibrosis, but tend to increase over time. (a) ETA concentrations of markers of inflammation, monocyte chemotaxis, and matrix remodeling are not associated with fibrosis at 24h. (b) Amphiregulin increases in patients but does significantly change over time in patients without fibrosis. MMP-9, S100A12, MMP-7, and CCL-2/MCP-1 levels increase over time in both groups, and are not significantly different between groups. TNF-α and P-selectin levels do not appear to change over time in either group. *p* values for individual timepoints are derived from logistic regression with log-transformed protein concentrations adjusted for age, sex, and enrollment hospital. *p* values for longitudinal analyses are derived from linear regression of the fold change between the later timepoint over the earlier timepoint and adjusted for age, sex, and enrollment hospital.

When steroid administration was added as a co-variate to the model, no changes were identified, and CCL-2/MCP-1 was associated with fibrosis at the 48-96h timepoint (**Supplemental Table 4**). A sensitivity analysis was also performed on these data, only analyzing measurements collected on patients with chest CT scans (**Supplemental Table 7**). CCL-2/MCP-1 was not significantly associated in patients at the 48-96h timepoint in this group, although this may have been due to study power (Est 1.16, [0.88-1.5], p=0.3 when adjusted for age, sex, and cohort). When presence of a chest CT was included as a co-variate in the model, the β for CT scan was 5.9 (p=0.01).

Again, we looked at how these markers changed over time, and did not find any significant pattern of change associated with fibrosis. Amphiregulin (FC 2.4, [0.8-6.7], p=0.09), MMP-9 (FC 2.7, [0.7-10.3], p=0.14), S100A12 (FC 5.6, [0.66-48], p=0.11), P-Selectin (FC 2.7, [0.6-11.6], p-0.18), MMP-7 (FC 1.7, [0.6-4.5], p=0.3), CCL-2/MCP-1 (FC 1.7, [0.3-9.2], p=0.5), and TNF-α (FC 2.7, [0.4-18), p=0.3) trended towards increasing over time in patients with fibrosis (**Figure 5b)**.

### There is only weak correlation between circulating and endobronchial fluid levels of measured proteins

We performed a correlation analysis between plasma and endotracheal aspirate measurements among patients who had paired samples at 24h (*n*= 30) **(Supplemental Figure 5).** We found that in general, plasma and endotracheal aspirate measurements of individual proteins were not positively correlated, and certain markers had slightly negative correlation (MMP-9, Pearson’s *r* = −0.19, and Amphiregulin, Pearson’s *r* = −0.13). However, some markers seemed to be correlated with one another, for example endotracheal aspirate P-selectin concentrations were positively correlated with IL-6 plasma levels (Pearson’s *r* = 0.65), CCL-13/MCP-4 plasma levels were correlated with MMP-9 endotracheal aspirate levels (Pearson’s *r* = 0.42), and MMP-9 plasma levels were positively correlated with CCL-2/MCP-1 endotracheal aspirate levels (Pearson’s *r* = 0.38).

## Discussion

In this study, we have identified protein biomarkers of radiographic fibrosis in patients who are critically ill with COVID-19. We found that a large proportion of patients with hypoxemic respiratory failure due to COVID-19 develop radiographic features of fibrosis (39 patients or 33% of cohort). These patients had a higher illness severity based on mechanical ventilation at time of ICU admission, enrollment APACHE score, and presence of ARDS. These patients also had a higher mortality and fewer ventilator-free days. These are clinical features that have previously been described as potentially contributing to the presence of fibroproliferative ARDS and post-ARDS fibrosis^2,4,5,23^.

A distinguishing feature of our study is the systematic review of chest imaging during hospitalization and the measurement of protein mediators of inflammation, monocyte chemotaxis, and extracellular matrix remodeling in plasma and ETA samples at multiple timepoints. The patients in our cohort who developed fibrosis had higher clinical indices of illness severity at ICU admission. However, when we looked at markers of general inflammation in the plasma and endotracheal aspirates of these patients, we did not find evidence of a “cytokine storm” or overwhelming inflammation. CRP levels have been used in mortality prediction models for COVID-19 respiratory failure^40,41^, but in our study CRP was not different between the two groups, suggesting that it is not only the inflammatory proteins that drive the development of fibrosis.

In contrast, markers of monocyte chemotaxis are elevated early in the plasma of patients with fibrosis. CCL-2/MCP-1 may be involved in monocyte chemotaxis to the alveolar space, leading to an imbalance of pro-fibrotic and pro-resolution macrophage populations^18,22,42–46^. CCL-13/MCP-4 has not previously been associated with ARDS illness severity or the development of pulmonary fibrosis, and further studies are needed to determine the significance of this novel association. It is clear that monocytes and macrophages play a role in the development of fibrotic remodeling and pulmonary fibrosis^22,46^. In COVID-19, dysregulated immune responses are implicated in severe disease. Our findings support prior conclusions that CCL2-expressing macrophages are found in the BAL of patients with COVID-19 ARDS,^18^ and a possible mechanism for ongoing injury is that resident macrophages secrete CCL2, and infiltrating monocytes that express alarmins (such as S100A12), inflammatory cytokines, and TGF-β enter the damaged space and differentiate into recruited alveolar macrophages, setting up a feed-forward loop of damage and fibrotic remodeling.

We had hypothesized that proteins previously shown to be elevated in patients with pulmonary fibrosis (Amphiregulin^32,35^, MMP-9^36,37^, MMP-7^37^, P-selectin^38^, S100A12^37^, and SDF-1a/CXCL12^39^) would be elevated in patients who develop fibrosis. Levels of TGF-β1, a marker strongly associated with fibrosis^36^, were also measured but the assay failed to meet quality control (**Supplemental Table 6**). We show that of the putative markers of fibrosis, Amphiregulin and MMP-9 are elevated in the plasma soon after ICU admission in patients with fibrosis. This suggests that there may be key distinctions between patients who go on to develop fibrosis and those who do not, and that these distinctions can be identified early in ICU course.

An advantage of our study is a longitudinal analysis of proteins of interest. When we look at paired samples over time, an increase in MMP-9 in the plasma may be protective against fibrosis, which is contrary to what we expected. This is opposite to the trend seen in ETAs, where MMP-9 levels increase over time in patients with fibrosis. MMP-9 is one of the measured proteins that was negatively correlated between plasma and ETA (**Supplemental Figure 5**). It is possible that ongoing fibrotic remodeling in the alveolar space is not reflected in the plasma along the same timescale of our measurements. Further studies with additional paired timepoints are necessary to identify how MMP-9 varies over time in both compartments in these patients. Further work is necessary to validate these findings in the context of other clinical patient cohorts. Many published ICU COVID-19 cohorts similar to ours do not have in-depth radiographic reports, which limits the ability to validate our findings.^47–51^ These markers should also be measured in patients convalescing from COVID-19, in particular those who have prolonged respiratory symptoms to understand how the levels of these proteins change in the plasma over time.

There are several limitations to our study. Our study was performed within one geographic region, but patients were enrolled in three area hospitals with different patient demographics and ICU practice patterns which can improve the generalizability of these findings. Within our initial cohort, patients with fibrosis were more likely to have a chest CT that was used to determine presence of fibrosis than patients without fibrosis (85% vs 39%, p<0.0001). This may have led to patients with fibrosis being classified as not having fibrosis, although this would likely bias our results towards the null hypothesis. When we performed a sensitivity analysis on patients with chest CTs, our results were no different in the plasma proteins, strengthening our findings. However, in the ETA, CCL-2/MCP-1 was no longer associated with fibrosis at the 48-96h timepoint. In addition, there was a significant discrepancy between patients with/without fibrosis in the interval of time between SARS-CoV-2 test positivity and image used to classify the patient into fibrosis/no fibrosis (**Supplemental Figure 2c**). This most likely reflects that patients who developed fibrosis had longer hospital stays (**Supplemental Figure 2b**) and images used to determine fibrosis were the closest to discharge or in-hospital death. Patients with fibrosis also had a longer time between SARS-CoV-2 test positivity and study enrollment (**Table 1**), because a higher proportion of these patients were transferred from other hospitals to our institution for specialized ARDS care. This could establish a lead time bias, in which patients with fibrosis have higher levels of these proteins because they had COVID-19 for longer prior to sampling. However, if this was the case, it would likely result in an underestimation of differences. Finally, it is unclear how many of the patients with radiographic features of fibrosis during hospitalization will go on to have persistent fibrotic changes. This is an active area of investigation in the field.

Our study defines a cohort of patients with COVID-19 who develop radiographic features of fibrosis during illness, and compares them to patients who did not develop these features despite critical illness and severe hypoxemic respiratory failure. Importantly, we show that differences in plasma proteins of monocyte chemotaxis and tissue remodeling occurred early following ICU enrollment, suggesting that there is a difference in these patients that exists before the development of fibroproliferative ARDS. Further, prolonged mechanical ventilation may not impact measurements at this early timepoint, separating the role of ventilator induced lung injury and viral induced lung injury. There may be individual immune host factors that increase the likelihood that a patient will develop fibrotic remodeling after injury. Further studies are necessary to identify the pathways that are involved. Our findings have implication for the development of biomarkers that may predict patients at risk for developing pulmonary fibrosis following hypoxemic respiratory failure, as well as elucidating potential pathways that could be targeted to improve outcomes of patients with viral induced ARDS.

## Methods

### Study Design

This is a prospective cohort study nested within a larger study that has been previously described^29,52^. We describe a cohort of patients admitted to three hospitals in Seattle, WA between March 16, 2020 and May 16, 2021 with clinical suspicion for COVID-19. Patients were excluded if they tested negative for COVID-19, did not have hypoxemic respiratory failure as defined by oxygen supplementation of high flow nasal cannula, noninvasive ventilation, or invasive mechanical ventilation at the time of enrollment, were not alive 10 days after enrollment, did not have chest imaging performed between day 3 of hospitalization and discharge or death, had pre-existing interstitial lung disease, age ≤ 18, pregnancy, or current incarceration. The median time to first chest CT was 10 days within the cohort, and therefore patients needed to be alive and in the hospital 10 days after enrollment to be included. This cutoff removes patients who died or were discharged before fibrosis could be identified. Some patients were admitted to the ICU for monitoring but did not have hypoxemic respiratory failure, and so only patients who were on noninvasive ventilation, high flow nasal cannula, or invasive mechanical ventilation were included in the analysis (*n*=151). Of these, 3 patients were excluded for pre-existing ILD, and the remaining 119 patients had chest imaging between day 3 and discharge or death.

All subjects were enrolled and had plasma collected within 24 hours of enrollment. A subset of patients had plasma collected approximately 7 days after ICU admission. Patients who were intubated had endotracheal aspirates collected within 24h of ICU admission and again approximately 48-96 hours after ICU admission. SARS-CoV-2 positive subjects were classified based on a positive SARS-CoV-2 RT-PCR nasal swab clinical test. Subjects were enrolled under a waiver of consent which was approved and supervised by the University of Washington IRB (Human Subjects Division Study: 9763). ETAs were also collected from critically ill patients with SARS-CoV-2 supported on invasive mechanical ventilation from Virginia Mason Franciscan Health Hospital (Benaroya Research Institute IRB number: 20-036). Patients/legal representatives were consented for data usage; if consent was withdrawn, patient data and samples were removed from the study. Some authors had access to information that could identify individual participants during data collection; this data was stored in a secure online database (RedCap).

### Analysis of CT scans

All patients had chest x-rays or chest CTs done between day 3 and discharge/death. These images were ordered as part of routine clinical care, and were not pre-specified. Images obtained closest to discharge or death were reviewed by a blinded chest radiologist for the presence of fibrosis. The chest CT scans were analyzed according to predominant pattern (ground glass opacities, consolidation, linear densities, reticulation, honeycombing, traction bronchiectasis, cysts, pneumatoceles), distribution (craniocaudal, axial, anterior/posterior), and overall disease extent (none, <5%, >5%). Finally, the pattern was identified as either fibrotic or non-fibrotic. For chest x-rays, predominant features of traction bronchiectasis were classified as a fibrotic pattern. Other features of fibrosis were not identified on chest x-rays. For patients enrolled from Virginia Mason Hospital (*n* = 17), only radiologist reports abstracted from the electronic medical record were used for analysis. Radiology reports that indicated “fibrosis”, “reticulation”, or “traction bronchiectasis” were classified as having fibrosis.

### Plasma and Endotracheal Aspirate Cytokine/Chemokine measurements

Cytokines and chemokines were measured from blood collected in an EDTA tube within 24 hours of study enrollment. A subset of patients had an additional blood sample collected between 7 days after study enrollment. The proteins were measured using electrochemiluminescent immunoassays per the manufacturer’s instructions ((V-Plex Proinflammatory Panel 1 (K15049D); V-Plex Chemokine Panel 1 (K15047D); V-Plex Cytokine Panel 1 (K15050D)). Mesoscale discovery U-Plex kits were used for total MMP-9, MMP-7, SDF-1a, TGF-β1, S100A12, and P-selectin. These plasma samples were collected in a sodium citrate tube but otherwise were processed the same way as those previously described. All plasma samples underwent two freeze-thaw cycles prior to analysis. Analytes that did not meet any of the following quality control parameters were excluded from subsequent analysis: 1) intraplate % CV > 25%; 2) interplate % CV > 25%; or 3) > 10% of samples with a measurement below the lower limit of detection.

ETAs were obtained by suctioning the endotracheal tube after instilling 10 mL of normal saline. The collected aspirate fluid was immediately mixed with an equal volume of 0.1% dithiothreitol and then placed on ice for 15 minutes to promote sample homogenization. Samples were then filtered through a 70 μm cell-strainer by gravity and flow-through was centrifuged at 400 x *g* for 10 minutes. The flow-through was immediately aliquoted and stored at −80°C until use. The samples underwent two freeze-thaw cycles prior to immunoassay analysis. We applied the V-Plex Pro-inflammatory (K15049D), V-Plex chemokine (K15047D), and U-Plex immunoassays on ETA samples as described above.

## Quantification and Statistical Analyses

### Outcome Definitions

We abstracted clinical data from the electronic medical record into standardized case report forms. ARDS was defined by the 2012 Berlin definition and chest x-rays were adjudicated for ARDS by a board-certified radiologist blinded to the primary data. APACHE III score was calculated based on the original instrument^53^. VFDs were defined as the total number of days alive and free of invasive mechanical ventilation in the 28 days following ICU admission^54^. Patients who died prior to day 28 were considered to have zero VFDs. RALE score was calculated as previously described^55^.

### Statistical Analyses

Our primary analysis tested for association between plasma or ETA protein concentrations and the primary outcome of radiographic features of fibrosis. We used multivariable regression and adjusted the log_2_-transformed protein concentration for age, sex, and APACHE III score (plasma) or admission hospital (ETA). To analyze how protein concentrations changed over time, we took the ratio of log transformed protein concentration at the later timepoint divided by the earlier timepoint, adjusted this for age, sex, and APACHE III (plasma) or admission hospital (ETA) and used linear regression to associate this with radiographic features of fibrosis. Additional analyses used steroids as a covariate in addition to age/sex/admission hospital.

For sensitivity analysis, use of CT scan to determine fibrosis status was added as a covariate to age, sex, and APACHE III score to obtain β values. Then, only patients with CT scans were selected for a statistical analysis. These results were compared to the entire cohort for concordance.

All analyses were performed in R version 4.1.1 and graphs were created in GraphPad Prism version 8.4.3.

## Declarations

### Ethical Approval

Subjects were enrolled under an emergency waiver of consent which was approved and supervised by the University of Washington IRB (Human Subjects Division Study: 9763). ETAs collected from critically ill patients with SARS-CoV-2 supported on invasive mechanical ventilation from Virginia Mason Franciscan Health Hospital were enrolled under a waiver of consent which was approved and supervised by the Benaroya Research Institute IRB (IRB number: 20-036).

### Author’s contributions

S.E.H. and C.M. designed the study and wrote the main manuscript text. S.E.H. performed data analysis and prepared figures. M.M. and S.E.H. ran assays. S.P. analyzed chest images. E.D.M, P.K.B., and M.M.W closely edited the manuscript and provided comments.

All authors reviewed the manuscript.

### Availability of data and materials

The datasets supporting the conclusions of this article are available from the authors on reasonable request.

## Data Availability

All data produced in the present study are available upon reasonable request to the authors

## Acknowledgements

The authors thank Sharon Sahi, Carolyn Brager, Sana S. Sakr, Neall Koetje, Ashley Garay, Brian Lee, Leslie Lazar, Sonya Homami, Grigory Loginov, Jana Zahlan, Hana Morris, Jada Roth and the Benaroya Research Institute COVID-19 Research Team for sample collection and processing. The authors also thank the patients, families, surrogates, and hospital clinical staff who contributed to this work during the COVID-19 pandemic.

