## Supplemental_Figures for "Mediators of monocyte chemotaxis and matrix remodeling are associated with the development of fibrosis in patients with COVID-19"

### Supplemental Figure 1

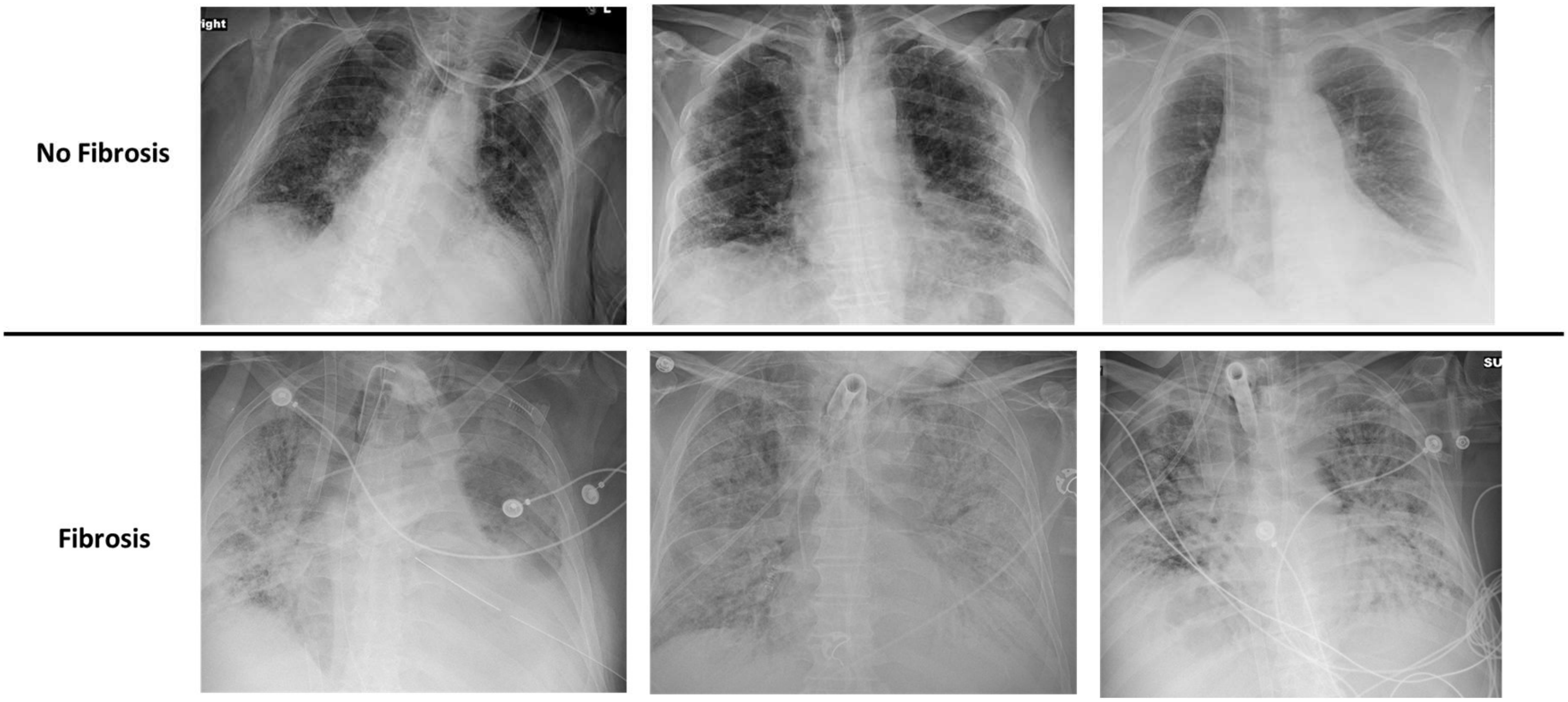

**Supplemental Figure 1: Representative chest x-rays from patients with fibrotic features (bottom row) and those with no evidence of fibrosis (top row).** Patients with fibrotic features had severe traction bronchiectasis, associated with severe ARDS

### Supplemental Figure 2

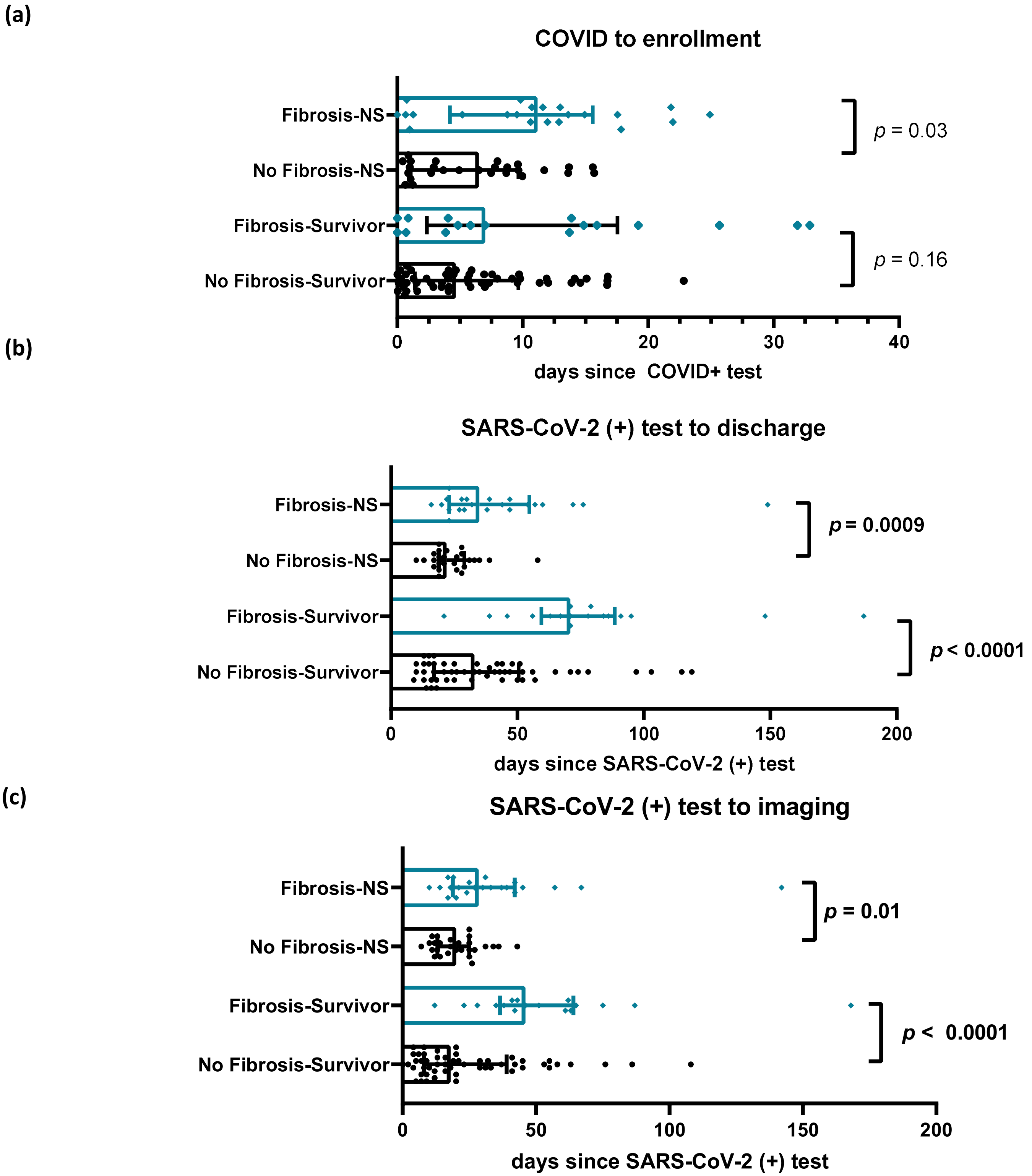

**Supplemental Figure 2: Time intervals between COVID+ PCR test and discharge, image used to classify fibrosis status, and study enrollment.** (a) Patients with fibrosis had a longer time between COVID+ test and study enrollment, likely due to an increased number of outside hospital transfers in this group. (b) Patients with radiographic evidence of fibrosis had longer hospital stays, this is true of both survivors and non-survivors (NS). (c) In general, patients with fibrosis had a longer time interval between positive COVID test and imaging used to determine fibrosis status (CXR or chest CT). *p* values reflect two-tailed Mann-Whitney tests of distribution. Survivor = survived to hospital discharge. NS = did not survive index hospital stay.

**Supplemental Figure 3**

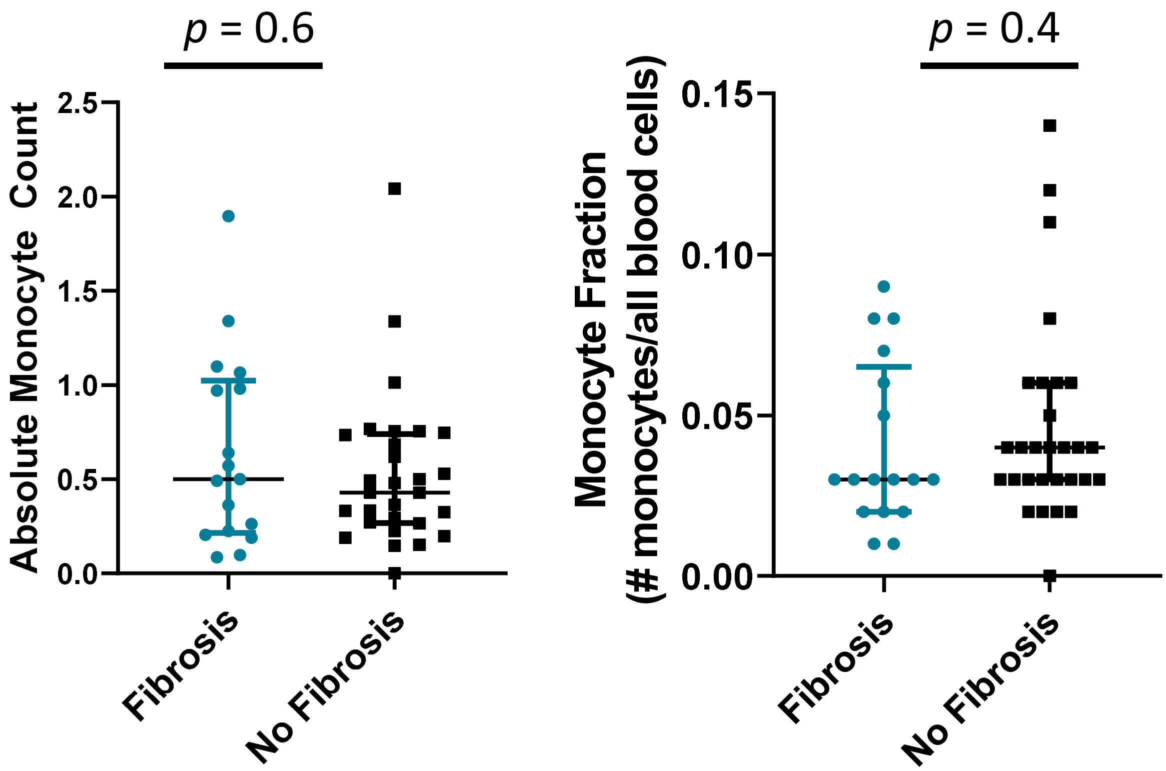

**Supplemental Figure 3: Monocyte levels are no different between patients with and without fibrosis 24h after study enrollment.** (a) Absolute monocyte count 24h after study enrollment (b) Monocyte fraction 24h after study enrollment (number of monocytes over all white blood cells). Cell counts were based on hospital laboratory complete blood cell count with automated differential. *p* values reflect Mann-Whitney tests.

**Supplemental Figure 4**

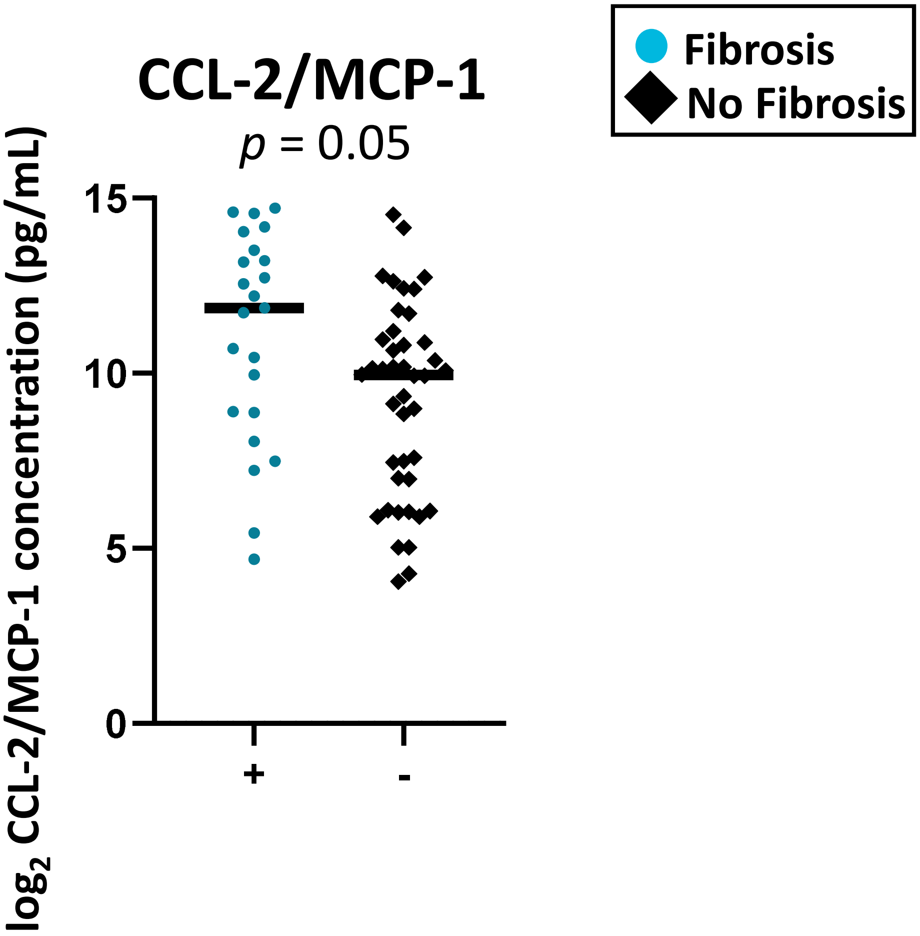

**Supplemental Figure 4: At 48-96h after study enrollment, ETA CCL-2/MCP-1 levels are higher in patients with fibrosis but not those without.** *p* value is derived from logistic regression with $log_2$ transformed protein concentration adjusted for age, sex, and enrollment hospital.

**Supplemental Figure 5**

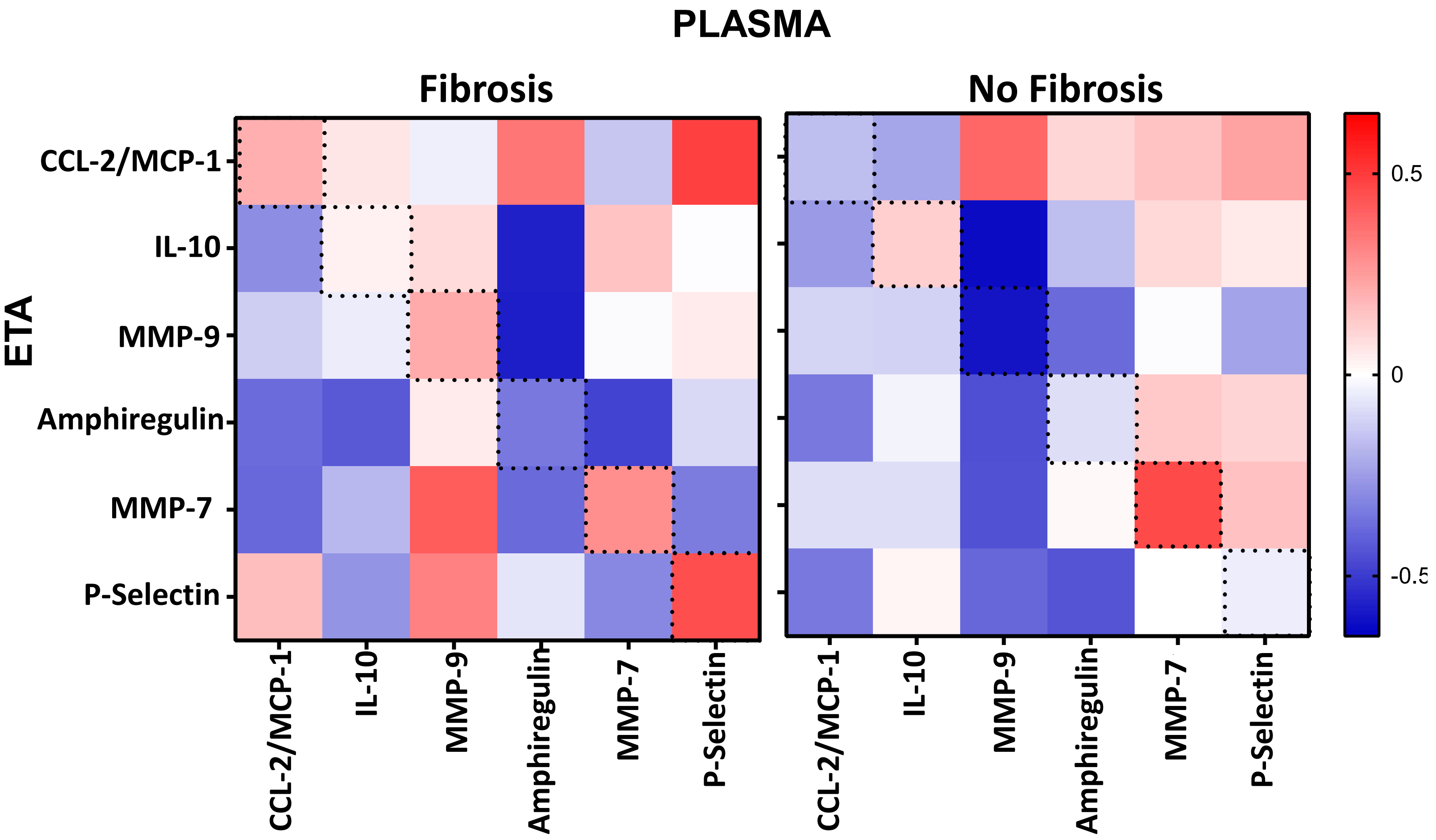

**Supplemental Figure 5: There is minimal correlation between plasma and ETA concentrations of proteins at 24h in patients with paired samples (Fibrosis n=19, No Fibrosis n = 18)**. Patients who developed fibrosis seemed to have higher correlation between plasma and ETA concentrations than patients without fibrosis. Spearman correlation was used to generate values. Outlined boxes represent the intersection of paired marker tests.

**Supplemental Table 1**

**Supplemental Table 1: Summary of chest CT features analyzed by chest radiologist.** Predominant abnormalities were summarized and reviewed by a chest radiologist, who gave a determination of “fibrosis” or “no fibrosis”.

| | Fibrosis (n=25) | No Fibrosis (n=7) |
|---|---|---|
| ***Predominant Abnormality*** | | |
| Ground Glass Opacities | 10 | 3 |
| Consolidation | 11 | 2 |
| Linear Densities | 1 | 2 |
| Reticulation | 3 | 0 |
| ***Reticulation*** | | |
| None | 10 | 7 |
| <5% | 5 | 0 |
| >5% | 10 | 0 |
| ***Honeycombing*** | | |
| None | 17 | 7 |
| <5% | 5 | 0 |
| >5% | 3 | 0 |
| ***Traction bronchiectasis*** | | |
| None | 9 | 6 |
| <5% | 5 | 1 |
| >5% | 11 | 0 |
| ***Imaging phenotype*** | | |
| Peripheral/Diffuse | 5 | 1 |
| Fibroproliferative ARDS | 12 | 0 |
| Unclassified fibrotic | 2 | 0 |
| Diffuse alveolar damage | 2 | 0 |
| Nonspecific | 4 | 6 |

### Supplemental Table 2

**Supplemental Table 2: Summary of statistics for plasma.** v1= 24h timepoint, v2= 3-7d timepoint

| **Plasma** | **Association of marker with presence of fibrosis** | | | | | | | | | | | | | | | |
|---|---|---|---|---|---|---|---|---|---|---|---|---|---|---|---|---|
| | ***Unadjusted*** | | | | ***Age, Sex*** | | | | ***Age, Sex, Apache*** | | | | ***Age, Sex, Apache, Steroids*** | | | |
| Marker | OR | CI | | p-value | OR | CI | | p-value | OR | CI | | p-value | OR | CI | | p-value |
| **IFNg v1** | 1.11 | 0.96 | 1.28 | 0.16 | 1.11 | 0.96 | 1.28 | 0.16 | 1.10 | 0.95 | 1.28 | 0.19 | 1.10 | 0.95 | 1.28 | 0.19 |
| **IFNg v2** | 0.93 | 0.58 | 1.51 | 0.77 | 1.02 | 0.58 | 1.77 | 0.95 | 1.00 | 0.57 | 1.75 | 1.00 | 0.97 | 0.53 | 1.75 | 0.91 |
| **IL-6 v1** | 1.21 | 1.01 | 1.46 | 0.04 | 1.22 | 1.01 | 1.48 | 0.04 | 1.17 | 0.96 | 1.42 | 0.12 | 1.19 | 0.97 | 1.45 | 0.087 |
| **IL-6 v2** | 1.28 | 0.81 | 2.00 | 0.27 | 1.35 | 0.78 | 2.33 | 0.26 | 1.34 | 0.76 | 2.36 | 0.30 | 1.31 | 0.74 | 2.32 | 0.32 |
| **IL-10 v1** | 1.00 | 0.76 | 1.32 | 1.00 | 0.99 | 0.75 | 1.31 | 0.94 | 0.94 | 0.70 | 1.27 | 0.69 | 0.95 | 0.71 | 1.27 | 0.71 |
| **IL-10 v2** | 0.83 | 0.45 | 1.54 | 0.55 | 0.86 | 0.44 | 1.67 | 0.64 | 0.92 | 0.45 | 1.90 | 0.81 | 0.88 | 0.42 | 1.86 | 0.73 |
| **CRP v1** | 0.93 | 0.70 | 1.24 | 0.64 | 0.94 | 0.71 | 1.26 | 0.68 | 0.92 | 0.69 | 1.24 | 0.58 | 0.92 | 0.68 | 1.23 | 0.56 |
| **CRP v2** | 0.71 | 0.41 | 1.26 | 0.23 | 0.65 | 0.35 | 1.24 | 0.18 | 0.64 | 0.33 | 1.26 | 0.18 | 0.63 | 0.31 | 1.27 | 0.18 |
| **CCL-2/MCP-1 v1** | 1.51 | 1.09 | 2.10 | 0.01 | 1.57 | 1.11 | 2.21 | 0.01 | 1.45 | 1.02 | 2.07 | 0.04 | 1.48 | 1.04 | 2.11 | 0.03 |
| **CCL-2/MCP-1 v2** | 0.96 | 0.50 | 1.85 | 0.90 | 0.93 | 0.48 | 1.82 | 0.83 | 0.94 | 0.49 | 1.81 | 0.85 | 0.92 | 0.48 | 1.76 | 0.78 |
| **CCL-13/MCP-4 v1** | 2.76 | 1.24 | 6.15 | 0.01 | 2.79 | 1.25 | 6.24 | 0.01 | 2.60 | 1.15 | 5.86 | 0.02 | 2.77 | 1.21 | 6.34 | 0.016 |
| **CCL-13/MCP-4 v2** | 1.18 | 0.25 | 5.72 | 0.83 | 1.15 | 0.22 | 6.14 | 0.86 | 1.11 | 0.20 | 6.27 | 0.90 | 0.9 | 0.15 | 5.45 | 0.9 |
| **TNF-a v1** | 1.56 | 1.02 | 2.38 | 0.04 | 1.56 | 1.02 | 2.38 | 0.04 | 1.37 | 0.90 | 2.10 | 0.14 | 1.37 | 0.9 | 2.1 | 0.14 |
| **TNF-a v2** | 0.86 | 0.35 | 2.10 | 0.73 | 0.85 | 0.33 | 2.19 | 0.72 | 1.15 | 0.32 | 4.09 | 0.82 | 1.21 | 0.34 | 4.35 | 0.75 |
| **Amphiregulin v1** | 1.94 | 1.17 | 3.24 | 0.01 | 2.17 | 1.26 | 3.76 | 0.01 | 1.98 | 1.11 | 3.52 | 0.02 | 2.02 | 1.13 | 3.62 | 0.019 |
| **Amphiregulin v2** | 1.23 | 0.82 | 1.85 | 0.31 | 1.32 | 0.86 | 2.02 | 0.20 | 1.38 | 0.87 | 2.17 | 0.17 | 1.21 | 0.74 | 1.96 | 0.44 |
| **MMP-9 v1** | 1.65 | 1.11 | 2.15 | 0.01 | 1.57 | 1.12 | 2.19 | 0.01 | 1.49 | 1.06 | 2.10 | 0.02 | 1.55 | 1.09 | 2.2 | 0.015 |
| **MMP-9 v2** | 0.99 | 0.69 | 1.44 | 0.97 | 1.00 | 0.69 | 1.46 | 0.99 | 1.03 | 0.69 | 1.54 | 0.88 | 1.01 | 0.67 | 1.5 | 0.97 |
| **MMP-7 v1** | 1.64 | 1.05 | 2.57 | 0.03 | 1.64 | 1.05 | 2.57 | 0.03 | 1.50 | 0.93 | 2.41 | 0.09 | 1.52 | 0.94 | 2.46 | 0.09 |
| **MMP-7 v2** | 1.63 | 1.11 | 2.39 | 0.01 | 1.62 | 1.09 | 2.39 | 0.02 | 1.58 | 1.02 | 2.45 | 0.04 | 1.68 | 1.07 | 2.64 | 0.03 |
| **P-Selectin v1** | 1.62 | 0.97 | 2.71 | 0.06 | 1.63 | 0.97 | 2.74 | 0.07 | 1.42 | 0.82 | 2.47 | 0.21 | 1.50 | 0.85 | 2.66 | 0.16 |
| **P-selectin v2** | 1.83 | 0.94 | 3.54 | 0.07 | 1.85 | 0.95 | 3.62 | 0.07 | 1.83 | 0.92 | 3.63 | 0.08 | 1.83 | 0.89 | 3.74 | 0.1 |
| **SDF-1a v1** | 1.48 | 0.88 | 2.49 | 0.13 | 1.51 | 0.89 | 2.55 | 0.12 | 1.34 | 0.78 | 2.31 | 0.28 | 1.41 | 0.81 | 2.47 | 0.22 |
| **SDF-1a v2** | 0.92 | 0.57 | 1.48 | 0.74 | 0.92 | 0.57 | 1.48 | 0.73 | 0.88 | 0.54 | 1.44 | 0.60 | 0.89 | 0.54 | 1.46 | 0.63 |

### Supplemental Table 3

**Supplemental Table 3: Plasma sensitivity analysis regarding use of chest CT for determining fibrosis status.** v1= 24h timepoint, v2= 3-7d timepoint. Logistic regression was used.

| | *Apache, Age, Sex covariates* | | | | *Patients with CT scans only, age, sex, apache covariates* | | | | *Patients without CT scans only, age, sex, apache covariates* | | | |
|---|---|---|---|---|---|---|---|---|---|---|---|---|
| Marker | OR | CI | | p-value | OR | CI | | p-value | OR | CI | | p-value |
| **IFNg v1** | 1.1 | 0.95 | 1.28 | 0.19 | 1.2 | 0.97 | 1.48 | 0.08 | 0.97 | 0.69 | 1.38 | 0.88 |
| **IFNg v2** | 1 | 0.57 | 1.75 | 1 | 0.44 | 0.01 | 16.74 | 0.52 | 0.91 | 0.21 | 4.01 | 0.89 |
| **IL-6 v1** | 1.17 | 0.96 | 1.42 | 0.12 | 1.17 | 0.91 | 1.51 | 0.23 | 1.47 | 0.77 | 2.83 | 0.24 |
| **IL-6 v2** | 1.34 | 0.76 | 2.36 | 0.3 | 0.79 | 0.24 | 2.62 | 0.57 | 4.54 | 0.02 | 1008.00 | 0.55 |
| **IL-10 v1** | 0.94 | 0.7 | 1.27 | 0.69 | 0.94 | 0.61 | 1.45 | 0.77 | 0.57 | 0.21 | 1.54 | 0.26 |
| **IL-10 v2** | 0.92 | 0.45 | 1.9 | 0.81 | | | | | | | | |
| **CRP v1** | 0.92 | 0.69 | 1.24 | 0.58 | 0.88 | 0.58 | 1.33 | 0.53 | 0.60 | 0.30 | 1.23 | 0.16 |
| **CRP v2** | 0.64 | 0.33 | 1.26 | 0.18 | | | | | | | | |
| **CCL-2/MCP-1 v1** | 1.45 | 1.02 | 2.07 | 0.04 | 1.48 | 0.92 | 2.38 | 0.1 | 1.03 | 0.45 | 2.34 | 0.95 |
| **CCL-2/MCP-1 v2** | 0.94 | 0.49 | 1.81 | 0.85 | | | | | 0.22 | 0.00 | 15.40 | 0.45 |
| **CCL-13/MCP-4 v1** | 2.6 | 1.15 | 5.86 | 0.02 | 3.32 | 1.06 | 10.4 | 0.04 | 1.69 | 0.20 | 14.46 | 0.62 |
| **CCL-13/MCP-4 v2** | 1.11 | 0.2 | 6.27 | 0.9 | | | | | | | | |
| **TNF-a v1** | 1.37 | 0.9 | 2.1 | 0.14 | 1.69 | 0.83 | 3.44 | 0.14 | 0.65 | 0.17 | 2.46 | 0.52 |
| **TNF-a v2** | 1.15 | 0.32 | 4.09 | 0.82 | | | | | | | | |
| **Amphiregulin v1** | 1.98 | 1.11 | 3.52 | 0.02 | 2.01 | 0.88 | 4.58 | 0.1 | 1.29 | 0.37 | 4.48 | 0.69 |
| **Amphiregulin v2** | 1.38 | 0.87 | 2.17 | 0.17 | 1.45 | 0.79 | 2.67 | 0.22 | 0.67 | 0.23 | 1.98 | 0.46 |
| **MMP-9 v1** | 1.49 | 1.06 | 2.1 | 0.02 | 1.56 | 1 | 2.43 | 0.05 | 1.83 | 0.50 | 6.69 | 0.35 |
| **MMP-9 v2** | 1.03 | 0.69 | 1.54 | 0.88 | 0.86 | 0.48 | 1.55 | 0.62 | 0.71 | 0.24 | 2.08 | 0.52 |
| **MMP-7 v1** | 1.5 | 0.93 | 2.41 | 0.09 | 1.42 | 0.72 | 2.81 | 0.31 | 0.59 | 0.17 | 2.12 | 0.41 |
| **MMP-7 v2** | 1.58 | 1.02 | 2.45 | 0.04 | 2.31 | 1.17 | 4.58 | 0.02 | 0.70 | 0.24 | 2.10 | 0.52 |
| **P-Selectin v1** | 1.42 | 0.82 | 2.47 | 0.21 | 1.11 | 0.54 | 2.25 | 0.78 | 3.16 | 0.71 | 14.06 | 0.13 |
| **P-selectin v2** | 1.83 | 0.92 | 3.63 | 0.08 | 1.93 | 0.79 | 4.72 | 0.15 | 1.27 | 0.22 | 7.38 | 0.78 |
| **SDF-1a v1** | 1.34 | 0.78 | 2.31 | 0.28 | 1.1 | 0.58 | 2.06 | 0.77 | 2.54 | 0.35 | 18.37 | 0.35 |
| **SDF-1a v2** | 0.88 | 0.54 | 1.44 | 0.6 | 0.99 | 0.54 | 1.82 | 0.97 | 0.66 | 0.25 | 1.76 | 0.40 |

### Supplemental Table 4

#### Supplemental Table 4. Clinical characteristics of combined cohort for ETA analysis

| Characteristics | No evidence of fibrosis (*n* = 47) | Evidence of fibrosis (*n* =27) |
|---|---|---|
| Age, average (std dev), range, yr | 50.8 (13.7), 20-77 | 51.6 (11.6), 21-72 |
| Male Sex, *n* (%) | 33 (70%) | 18 (67%) |
| Ethnicity (% Hispanic) | 19 (40%) | 12 (44%) |
| Race (% African American) | 3 (4%) | 0 |
| OSH Transfer, *n* (%) | 35 (74%) | 26 (96%) |
| ARDS at enrollment, *n* (%) | 40 (85%) | 25 (92%) |
| Received dexamethasone | 31 (66%) | 24 (89%) |
| APACHE, median (IQR) | 83 (65-98.8) | 96 (87.5-107) |
| In-hospital death, *n* (%) | 14 (30%) | 15 (56%) |
| VFDs, median (IQR), days | 4 (0-13.5) | 0 (0-0.5) |

### Supplemental Table 5

**Supplemental Table 5. Quality Control analysis for assays used for plasma and ETA measurements**. Data were not included in the analysis if 1) intraplate %CV >25%, 2) interplate %CV >25%, or 3) >10% of samples with a measurement below the lower limit of detection

**Biomarker Quality Control - Plasma**

| Biomarker | Out of range/imputed (%) | Average Intraplate % CV | Interplate %CV | Included in analysis |
|---|---|---|---|---|
| IL-6 | 1 | 8.71 | 4.65 | yes |
| IFN-γ | 6 | 8.1 | 4.8 | yes |
| TNF-α | 1 | 5.9 | 4.9 | yes |
| IL-10 | 0 | 6.1 | 4.7 | yes |
| CRP | 0 | 8.6 | 10.3 | yes |
| CCL-2/MCP-1 | 0 | 5.4 | 10.4 | yes |
| CCL-13/MCP-4 | 0 | 6 | 18.9 | yes |
| Amphiregulin | 0 | 7.6 | 23.8 | yes |
| MMP-9 | 0.3 | 4.8 | 15.2 | yes |
| MMP-7 | 0 | 4 | 7.5 | yes |
| P-Selectin | 0 | 3.4 | 7.2 | yes |
| S100A12 | 26 | 10.3 | 26.2 | no |
| SDF-1a/CXCL12 | 7.7 | 11.3 | 22.7 | yes |
| TGF-β1 | 2.5 | 22 | 37.3 | no |

**Biomarker Quality Control - ETA**

| Biomarker | Hospital | Out of range/imputed (%) | Intraplate % CV | Interplate %CV | Included in analysis |
|---|---|---|---|---|---|
| IL-6 | *HMC* | 0 | 20.8 | 48.7 | no |
| | *VM* | 8.6 | 8.7 | NA | yes |
| IFN-γ | *HMC* | 67 | 1.5 | 12.1 | no |
| | *VM* | 15 | 0.49 | NA | yes |
| TNF-α | *HMC* | 3.5 | 1.4 | 3.5 | yes |
| | *VM* | 2.8 | 7.3 | NA | yes |
| CCL2/MCP-1 | *HMC* | 2.5 | 3.2 | 14.9 | yes |
| | *VM* | 8.6 | 4.3 | NA | yes |
| CCL13/MCP-4 | *HMC* | 33 | 9.3 | 68.9 | no |
| | *VM* | 27 | 6.9 | NA | yes |
| Amphiregulin | | 0 | NA | 6.7 | yes |
| MMP-9 | | 0 | NA | 4.8 | yes |
| MMP-7 | | 3.5 | 2.2 | 5.9 | yes |
| P-Selectin | | 1.4 | NA | 12.5 | yes |
| S100A12 | | 9.7 | 6.7 | 7.1 | yes |
| SDF-1a/CXCL12 | | 52.6 | NA | 44 | no |
| TGF-β1 | | 68.4 | NA | NA | no |

### Supplemental Table 6

#### Supplemental Table 6. Summary of ETA statistics

v1= 24h timepoint, v2= 3-7d timepoint

**ETA data** — **Logistic regression. Association of $log_2$-transformed marker with presence of fibrosis**

| | ***Unadjusted*** | | | | ***Age, Sex*** | | | | ***Age, Sex, Cohort*** | | | | ***Age, Sex, Cohort, Steroids*** | | | |
|---|---|---|---|---|---|---|---|---|---|---|---|---|---|---|---|---|
| Marker | OR | CI | | p-value | OR | CI | | p-value | OR | CI | | p-value | OR | CI | | p-value |
| IL-10 v1 | 0.94 | 0.77 | 1.16 | 0.56 | 0.94 | 0.76 | 1.16 | 0.54 | 0.90 | 0.72 | 1.13 | 0.37 | 0.89 | 0.71 | 1.12 | 0.31 |
| IL-10 v2 | 1.11 | 0.96 | 1.28 | 0.17 | 1.11 | 0.96 | 1.29 | 0.16 | 1.05 | 0.89 | 1.24 | 0.58 | 1.05 | 0.89 | 1.24 | 0.57 |
| MCP-1 v1 | 1.15 | 0.90 | 1.47 | 0.27 | 1.22 | 0.92 | 1.62 | 0.15 | 1.26 | 0.94 | 1.69 | 0.12 | 1.26 | 0.94 | 1.7 | 0.12 |
| MCP-1 v2 | 1.27 | 1.04 | 1.55 | 0.02 | 1.27 | 1.04 | 1.56 | 0.02 | 1.22 | 1.00 | 1.50 | 0.05 | 1.23 | 1 | 1.51 | 0.05 |
| TNF-a v1 | 0.95 | 0.75 | 1.20 | 0.63 | 0.95 | 0.75 | 1.21 | 0.67 | 0.94 | 0.74 | 1.19 | 0.58 | 0.9 | 0.7 | 1.16 | 0.42 |
| TNF-a v2 | 1.09 | 0.92 | 1.30 | 0.32 | 1.10 | 0.92 | 1.31 | 0.30 | 0.99 | 0.81 | 1.22 | 0.96 | 1.0 | 0.82 | 1.26 | 0.89 |
| Amphiregulin v1 | 0.88 | 0.54 | 1.42 | 0.58 | 0.85 | 0.52 | 1.40 | 0.52 | 0.88 | 0.53 | 1.45 | 0.60 | 0.88 | 0.52 | 1.46 | 0.6 |
| Amphiregulin v2 | 1.24 | 0.86 | 1.78 | 0.25 | 1.22 | 0.83 | 1.80 | 0.30 | 1.23 | 0.83 | 1.83 | 0.29 | 1.27 | 0.84 | 1.91 | 0.25 |
| MMP-9 v1 | 0.90 | 0.69 | 1.18 | 0.44 | 0.90 | 0.68 | 1.18 | 0.42 | 0.86 | 0.64 | 1.16 | 0.31 | 0.84 | 0.62 | 1.15 | 0.27 |
| MMP-9 v2 | 1.01 | 0.80 | 1.28 | 0.94 | 1.00 | 0.78 | 1.28 | 0.99 | 0.96 | 0.74 | 1.25 | 0.78 | 0.99 | 0.75 | 1.3 | 0.9 |
| MMP-7 v1 | 1.06 | 0.73 | 1.54 | 0.74 | 1.03 | 0.70 | 1.52 | 0.86 | 1.04 | 0.70 | 1.54 | 0.83 | 1.2 | 0.7 | 1.5 | 0.9 |
| MMP-7 v2 | 1.14 | 0.82 | 1.57 | 0.43 | 1.14 | 0.80 | 1.61 | 0.46 | 1.16 | 0.81 | 1.65 | 0.42 | 1.2 | 0.8 | 1.8 | 0.32 |
| P-Selectin v1 | 1.07 | 0.80 | 1.44 | 0.62 | 1.06 | 0.79 | 1.43 | 0.68 | 1.09 | 0.81 | 1.48 | 0.56 | 1.1 | 0.8 | 1.5 | 0.5 |
| P-selectin v2 | 1.16 | 0.91 | 1.46 | 0.22 | 1.15 | 0.90 | 1.46 | 0.26 | 1.13 | 0.89 | 1.44 | 0.31 | 1.2 | 0.9 | 1.5 | 0.27 |
| S100A12 v1 | 0.97 | 0.85 | 1.11 | 0.70 | 0.97 | 0.85 | 1.11 | 0.67 | 0.97 | 0.84 | 1.11 | 0.61 | 0.97 | 0.84 | 1.1 | 0.63 |
| S100A12 v2 | 1.13 | 0.91 | 1.41 | 0.27 | 1.14 | 0.90 | 1.43 | 0.27 | 1.11 | 0.89 | 1.39 | 0.35 | 1.1 | 0.9 | 1.5 | 0.34 |

### Supplemental Table 7

**Supplemental Table 7. Sensitivity Analysis for ETA Statistics**
v1= 24h timepoint, v2= 3-7d timepoint

| **ETA data** | ***Age, Sex, Cohort*** | | | | ***Patients with CT scans only, age, sex, cohort covariates*** | | | | ***Patients without CT scans only, age, sex, cohort covariates*** | | | |
|---|---|---|---|---|---|---|---|---|---|---|---|---|
| Marker | OR | CI | | p-value | OR | CI | | p-value | OR | CI | | p-value |
| IL-10 v1 | 0.90 | 0.72 | 1.13 | 0.37 | 0.79 | 0.52 | 1.19 | 0.24 | 1.02 | 0.68 | 1.5 | 0.91 |
| IL-10 v2 | 1.05 | 0.89 | 1.24 | 0.58 | 0.95 | 0.76 | 1.2 | 0.67 | 1.27 | 0.86 | 1.9 | 0.22 |
| MCP-1 v1 | 1.26 | 0.94 | 1.69 | 0.12 | 1.43 | 0.86 | 2.4 | 0.16 | 1.26 | 0.67 | 2.3 | 0.45 |
| MCP-1 v2 | 1.22 | 1.00 | 1.50 | 0.05 | 1.16 | 0.88 | 1.5 | 0.27 | 1.17 | 0.77 | 1.78 | 0.44 |
| TNF-a v1 | 0.94 | 0.74 | 1.19 | 0.58 | 0.88 | 0.6 | 1.3 | 0.48 | 0.85 | 0.50 | 1.4 | 0.49 |
| TNF-a v2 | 0.99 | 0.81 | 1.22 | 0.96 | 0.88 | 0.64 | 1.2 | 0.42 | 1.13 | 0.76 | 1.67 | 0.52 |
| Amphiregulin v1 | 0.88 | 0.53 | 1.45 | 0.60 | 0.74 | 0.31 | 1.7 | 0.47 | 0.79 | 0.27 | 2.3 | 0.63 |
| Amphiregulin v2 | 1.23 | 0.83 | 1.83 | 0.29 | 1.15 | 0.61 | 2.17 | 0.65 | 1.52 | 0.74 | 3.1 | 0.24 |
| MMP-9 v1 | 0.86 | 0.64 | 1.16 | 0.31 | 0.73 | 0.42 | 1.3 | 0.26 | 0.86 | 0.50 | 1.5 | 0.57 |
| MMP-9 v2 | 0.96 | 0.74 | 1.25 | 0.78 | 0.9 | 0.64 | 1.25 | 0.51 | 0.97 | 0.51 | 1.8 | 0.93 |
| MMP-7 v1 | 1.04 | 0.70 | 1.54 | 0.83 | 0.98 | 0.5 | 1.9 | 0.9 | 0.86 | 0.43 | 1.7 | 0.65 |
| MMP-7 v2 | 1.16 | 0.81 | 1.65 | 0.42 | 1.05 | 0.6 | 1.7 | 0.86 | 1.29 | 0.61 | 2.7 | 0.49 |
| P-Selectin v1 | 1.09 | 0.81 | 1.48 | 0.56 | 0.9 | 0.55 | 1.48 | 0.67 | 1.24 | 0.54 | 2.8 | 0.58 |
| P-selectin v2 | 1.13 | 0.89 | 1.44 | 0.31 | 1.1 | 0.75 | 1.5 | 0.73 | 1.24 | 0.54 | 2.8 | 0.58 |
| S100A12 v1 | 0.97 | 0.84 | 1.11 | 0.61 | 0.81 | 0.5 | 1.3 | 0.35 | 1.03 | 0.79 | 1.35 | 0.81 |
| S100A12 v2 | 1.11 | 0.89 | 1.39 | 0.35 | 0.97 | 0.7 | 1.35 | 0.84 | 1.08 | 0.72 | 1.6 | 0.70 |
